# Fluorescent Antibody-Based Detection and Ultrastructural Analysis of *Streptococcus pneumoniae* in Human Sputum

**DOI:** 10.1101/2024.10.22.24314814

**Authors:** Ana G. Jop Vidal, Meg Francis, Maneesha Chitanvis, Ithiel J. Frame, Poonam Sharma, Patricio Vidal, Claudio F. Lanata, Carlos Grijalva, William Daley, Jorge E. Vidal

**Affiliations:** Department of Cell and Molecular Biology; Center for Immunology and Microbial Research; Department of Pathology, University of Mississippi Medical Center, Jackson MS; Rollins School of Public Health; Emory University, Atlanta GA; Instituto de Investigacion Nutricional; Lima, Peru; Department of Health Policy, Vanderbilt University Medical Center, Nashville, TN

## Abstract

**Background:** Pneumococcal pneumonia continues to be a significant global health burden, affecting both children and adults. Traditional diagnostic methods for sputum analysis remain challenging. The objective of this study was twofold: to develop a rapid and easy-to-perform assay for the identification of *Streptococcus pneumoniae* (Spn) directly in sputum specimens using fluorescence microscopy, and to characterize with high-resolution confocal microscopy the ultrastructure of pneumococci residing in human sputum.

**Methods:** We fluorescently labeled antibodies against the pneumococcal capsule (Spn-FLUO). The specificity and sensitivity of Spn-FLUO for detecting Spn was evaluated *in vitro* and *in vivo* using mouse models of carriage and disease, human nasopharyngeal specimens, and sputum from patients with pneumococcal pneumonia. Spn was confirmed in the specimens using culture and a species-specific qPCR assays. Confocal microscopy and Imaris software analysis were utilized to resolve the ultrastructure of pneumococci in human sputum.

**Results:** Compared with cultures and qPCR, Spn-FLUO demonstrated high sensitivity (78-96%) in nasopharyngeal samples from mice and humans. The limit of detection (LOD) in nasopharyngeal samples was ≥1.6×10⁴ GenEq/ml. The specificity in human nasopharyngeal specimens was 100%. In lung specimens from mice infected with pneumococci, Spn-FLUO reached 100% sensitivity with a LOD of ≥1.39×10⁴ GenEq/ml. In human sputum, the sensitivity for detecting Spn was 92.7% with a LOD of 3.6×10³ GenEq/ml. Ultrastructural studies revealed that pneumococci are expectorated as large aggregates with a median size of 1336 µm².

**Conclusions:** Spn-FLUO is a rapid and sensitive assay for detecting Spn in human sputum within 30 min. The study highlights that most pneumococci form aggregates in human sputum.

## Background

*Streptococcus pneumoniae* (Spn) strains infect the lungs causing pneumococcal pneumonia, a common community-acquired pneumonia (CAP) that causes over a million deaths annually worldwide^1^. Pneumococcal pneumonia is an acute bacterial infection of the pulmonary parenchyma caused by colonization of the lung and subsequent, or concurrent, invasion of pneumococci into the respiratory epithelium^2, 3^. Invasion of bronchi and terminal bronchiole leads pneumococci to reach respiratory bronchiole, alveolar ducts, and alveoli, where pneumococci have access to the alveolar-capillary network, a large surface area containing ∼300 billion of blood capillaries and where the exchange of CO_2_ by O_2_ occurs.

Spn strains encompass over 100 capsular types; the capsule is a target of pneumococcal vaccines that have saved millions since their licensure^4^. The capsular polysaccharide is negatively charged, enabling pneumococci to colonize the lungs by evading entrapment by mucus secreted in the lumen of bronchi and terminal bronchiole^2^. The capsule also acts as an anti-phagocytic factor by blocking complement deposition^2, 5, 6, 7^. Consequently, expression of the pneumococcal capsule is maximized during pneumococcal pneumonia and it was utilized in this study to identify the presence of Spn in clinical specimens.

The diagnosis of pneumococcal pneumonia is based on symptoms and suggestive radiographic findings^8, 9^. Diagnostic testing to determine pneumococcal pneumonia is optional for non-hospitalized patients, while those requiring admission should undergo blood cultures, respiratory cultures, and Gram staining of good-quality sputum. If the disease is severe, a urinary antigen test may also be considered^8^. However, cultures have poor sensitivity and can take at least 24 h to yield results. Moreover, the urinary antigen test for identifying pneumococcal pneumonia has limited sensitivity and specificity^8^.

Pneumonia multiplex (m)PCR panels have demonstrated superior performance in detecting respiratory pathogens, including pneumococcal strains. mPCR panels offer improved specificity and turnaround time compared to traditional methods^10^. Other molecular assays include real-time PCR methods targeting a number of species-specific genes, or gene sequences, such as the gene encoding the autolysin *lytA*^11, 12^, the pneumolysin *ply*^12^ gene, the permease gene *piaB* of the iron transporter PiaABC^13^ or a putative transcriptional regulator SP2020^14^. These PCR assays typically exhibit high sensitivity and specificity (>90%), and depending on the platform, real-time assays can deliver results within 3-4 h.

Prompt and accurate identification of pneumococcal pneumonia as the etiology is crucial for tailoring antibiotic management to a specific patient, which is paramount to preventing complications and mortality^8, 15^. Early administration of antibiotic therapy has been demonstrated to improve short-term mortality rates in both human patients and animal models of pneumococcal disease^16, 17^. Antibiotic therapy for community-acquired pneumonia (CAP) is typically initiated empirically within 1-2 h of presentation to a healthcare facility, prior to the identification of the causative pathogen^16^. However, the indiscriminate use of empirically prescribed antibiotics has contributed to a significant increase in antimicrobial resistance rates, particularly in certain geographical regions. For example, in some areas, over 70% of Spn strains isolated from pneumococcal pneumonia cases are resistant to first-line macrolide antibiotics^18, 19^.

Addressing the global burden of antibiotic resistance requires multifaceted efforts, including ensuring equitable access to, and developing innovative, diagnostics. In this study, we fluorescently labeled antibodies against the pneumococcal capsule, hereafter referred to as Spn-FLUO, and evaluated their performance in detecting pneumococcal strains within 30 min. Given the capsule’s high expression during pneumococcal disease, we hypothesized that antibodies targeting the capsule would enhance the rapid detection of strains in lower airway specimens. We systematically tested Spn-FLUO to detect Spn strains grown *in vitro*, as well as strains colonizing the upper and lower airways of mice in murine models of carriage and disease. Additionally, we assessed the efficacy of Spn-FLUO in detecting Spn strains colonizing the nasopharynx of children. We evaluated the diagnostic performance of Spn-FLUO in identifying pneumococcal pneumonia as the etiology in specimens collected from patients with pneumococcal pneumonia. Results were obtained within 30 min of sample collection.

Given the distinctive characteristics of the pneumococci observed in the sputum, we conducted additional ultrastructural studies using high-resolution laser scanning confocal microscopy and advanced image analysis. Consequently, we present novel insights into the ultrastructure of pneumococcal aggregates formed in human sputum.

## Methods

### Bacterial strains, culture media, and antibiotics

Spn reference strains and mutants used in the present study were: TIGR4^20^, SPJV41^21, 22^, D39^23^, EF3030^24^, S19F 4924^25^ and S6B 8655^26^. Strains were routinely cultured on blood agar plates (BAP), or grown in Todd Hewitt broth containing 0.5% (w/v) yeast extract (THY), at 37°C with a 5% CO_2_ atmosphere. Where indicated, streptomycin (Str, 200 µg/ml), trimethoprim (Tmp, 10 µg/ml), tetracycline (Tet, 1 µg/ml), or/and erythromycin (Ery, 1 µg/ml) was added to BAP. All antibiotics were purchased from Millipore-Sigma Sigma-Aldrich.

### Preparation of inocula

The inoculum was prepared as previously described^27^. Briefly, bacteria were inoculated on blood agar plates (BAP) and incubated overnight at 37°C in a 5% CO_2_ atmosphere. Bacteria were then harvested from plates by PBS washes and this suspension was used to inoculate THY or cell cultures, that was brought to a final OD_600_ of ∼0.1. This suspension contained ∼5.15×10^8^ cfu/ml as verified by serial dilution and platting of aliquots of the suspension.

### Cell cultures

Human pharyngeal Detroit 562 cells (ATCC CCL-138) were cultured in Eagle’s minimum essential medium (MEM) (Gibco) supplemented with non-heat-inactivated 10% fetal bovine serum (FBS) from R&D systems, 1% nonessential amino acids (Gibco), 1% glutamine (Gibco), penicillin (100 U/ml), and streptomycin (100 µg/ml), and the pH was buffered with HEPES (10 mM) (Gibco). Cells were harvested with 0.25% trypsin (Gibco), resuspended in the cell culture medium, and incubated at 37°C in a 5% CO_2_ humidified atmosphere.

### Fluorescent labeling of anti-capsule antibodies

Antibodies were purchased from the Staten Serum Institute (Denmark). The following antibodies were used throughout this study: Omni antiserum (recognizes 92 serotypes), type serum 2, type serum 4, group serum 6, and group serum 19. Serum was depleted of proteins less than 100 kDa using an Amicon Ultra-100K centrifugal filter (Millipore-Sigma), washed three times with PBS. Proteins were quantified using Bradford reagent, and 1 mg of the antibodies was labeled with either Alexa-488 or Alexa-555 following the manufacturer’s recommendations (Molecular Probes). After purification by size exclusion chromatography (≥40 kDa), collected fractions were assessed for reactivity against the specific serotype, serogroup, or several serotypes for the Omni antiserum. Proteins were quantified from reactive fractions using the Bradford assay, and antisera were stored at 4°C for short-term or -20°C for long-term storage. Labeled antibodies stored at 4°C remained reactive for up to three months, while those frozen have been effective in labeling bacteria for over three years. For the sake of simplicity, the fluorescently labeled Omni antiserum is referred to as Spn-FLUO throughout this manuscript.

### *In vitro* imaging of pneumococci

Pneumococci were inoculated into 8-well glass slides (Lab-Tek^®^) and incubated for 4 h at 37°C in a 5% CO_2_ atmosphere. Some slides contained cultures of Detroit 562 cells (ATCC CCL-198) grown to confluence, typically 7 days post seeding. Pneumococci attached to the glass substratum or human cells were then washed twice with PBS and fixed with 2% PFA for 15 min at room temperature. After removing the 2% PFA, the bacteria were washed with PBS and blocked with 2% bovine serum albumin (BSA) for 1 h at 37°C. The pneumococci were then incubated with the specified labeled antibodies (∼20 μg/ml) for 1 h at room temperature. The stained preparations were subsequently washed twice with PBS and mounted with ProLong Diamond Antifade Mountant with DAPI (Molecular Probes). In some preparations, DAPI was replaced with TO-PRO-3 (1 μM), a carbocyanine monomer nucleic acid stain (Molecular Probes). Fluorescence images were acquired using an upright, epi-fluorescence Nikon Ni-U Research Microscope equipped with a Nikon DS-Qi2 sCMOS Camera system. Confocal images were obtained using a Nikon C2 laser scanning confocal microscopy system, and the micrographs were analyzed with ImageJ version 1.49k (National Institutes of Health, USA) or Imaris software (Bitplane).

### Mouse models of pneumococcal nasopharyngeal carriage and disease

Inbred 6-7 week-old C57BL/6 mice -pneumococcal carriage-, or 4-5 week-old Balb/c mice -pneumococcal pneumonia-model (Charles River Laboratories) were anesthetized with 5% isoflurane (vol/vol) in oxygen (2 liters/min) administered via an RC2 calibrated vaporizer (VetEquip Inc) and then infected with ∼1×10^5^ CFU of strain Spn EF3030 (carriage model) or ∼1×10^8^ cfu of TIGR4 or SPJV41 (pneumonia model). The animals were weighed daily, and their behavior and appearance were monitored twice daily for up to four days. Mice in the pneumonia model were euthanized when they lost ≥20% of their body weight, compared to their body weight before infection, or when mice were non-responsive to manual stimulation, and/or if they show signs of illness such as ruffled fur, intermittent hunching, and exhibiting respiratory distress. Mice were then euthanized, and the upper airways, including nasopharyngeal tissue or the lungs, were aseptically collected, placed in THY with 10% glycerol and immediately homogenized. For fluorescence staining (explained below), the nasopharynx from some mice was collected and fixed with 10% PFA. The specimens were then stored at -80°C. This aliquot was used for bacterial counts and detection with labeled antibodies. To obtain bacterial counts, the homogenized tissue was serially diluted in PBS and plated onto BAP with gentamicin. The Institutional Animal Care and Use Committee (IACUC) at the University of Mississippi Medical Center approved the protocol used in this study (1584), overseeing the welfare, well-being, and proper care of all mice utilized in this study. All mouse experiments followed the guidelines outlined in the National Science Foundation Animal Welfare Act (AWA).

### Fluorescence staining of nasopharyngeal tissue

The PFA-fixed nasopharynxes were paraffin-embedded, sectioned (∼5 µm), mounted on a slide and deparaffinized. Tissues were washed three times with PBS and the preparations were blocked with 2% bovine serum albumin (BSA) for 1 h at room temperature. These preparations were then incubated for 1 h with serogroup-specific (S19) polyclonal antibody (Statens Serum Institute, Denmark) (20 μg/ml) that had been previously labeled with Alexa-555 (Molecular Probes). Stained tissues were finally washed three times with PBS, air dried and then mounted with ProLong Diamond Antifade mounting medium containing DAPI (Molecular Probes). Confocal micrographs were obtained using a Nikon C2 laser scanning confocal microscopy system and analyzed using the Imaris software (Bitplane).

### Specimens from Humans with Pneumococcal Pneumonia

Human specimens, including sputum and bronchoalveolar lavages from de-identified patients with microbiologically and radiologically confirmed pneumococcal pneumonia, were utilized in this study. Institutional Review Board (IRB) exemption was obtained for the use of these de-identified specimens. When available, the age of the patients ranged from 19 months through 60 years. The specimens were collected according to institutional guidelines by the University of Mississippi Medical Center’s clinical laboratory and stored at -80°C until processed. As part of the microbiological diagnostics, the pathology laboratory at UMMC isolated a Spn strain from each specimen. Additionally, the Spn etiology was confirmed by *lytA*-based real-time PCR, as detailed^28, 29^ and briefly described below.

### Human nasopharyngeal specimens

Human nasopharyngeal specimens (N=50) utilized in the current study had been collected from Peruvian children and processed for pneumococcal detection and quantification by qPCR and culture in previous studies^30, 31^. Children enrolled in the mentioned studies were aged 0-3 years of age; details on the study population have been published elsewhere^31, 32, 33^. De-identified specimens were processed at Emory University, obtaining IRB exemption. Briefly, these nasopharyngeal samples were collected following WHO recommendations^34^ with a deep NP swab, using rayon polyester swabs and were immediately placed in 2.0 ml cryogenic vials with 1 ml of transport medium, a mixture containing skim milk-tryptone-glucose-glycerin (STGG) and then stored at -80°C^35^.

### Identification of pneumococcus using Spn-FLUO in mouse tissue or human specimens

Specimens were thawed on ice and an aliquot (10 µl) of each specimen was immediately mixed with Spn-FLUO (50 μg/ml) and incubated for 10 min at room temperature in the dark. Five µl of this suspension was dropped onto a microscope slide and covered with a coverslip. The stained sample was analyzed within five minutes using an upright, epi-fluorescence Nikon Ni-U Research Microscope equipped with a Nikon DS-Qi2 sCMOS Camera system. We first situated the specimen in the correct optical plane (i.e., visualizing cells, tissue, and/or bacteria) with the 60x objective and a bright light source. The slides were then analyzed and scored using epifluorescence in the green channel, following a semiquantitative algorithm: 1-5 pneumococci per field in at least three fields corresponded to “+”, 5-10 pneumococci per field to “++”, and more than 10 pneumococci per field was scored as “+++”. Negative samples had no pneumococci observed. Micrographs from positive specimens were obtained using the 60x and 100x objectives.

### Staining and confocal analysis of human sputum

An aliquot of sputum sample (10 µl) was dropped onto a slide and air dry for ∼20 min at room temperature. The sputum was then stained for 1 h with Spn-FLUO (20 μg/ml) and wheat germ agglutinin conjugated to Alexa-555 [(WGA), 5 µg/ml]. The stained specimen was washed three times with PBS, air-dried, and mounted with ProLong Diamond Antifade mounting medium containing DAPI (Molecular Probes). Confocal micrographs were obtained using a Nikon C2 laser scanning confocal microscopy system and z-stacks micrographs were analyzed using the NIS Elements Basic Research software, version 4.30.01 build 1021. For 3D visualization, creation of animations, and quantification purposes, images were processed using the Imaris software 64x Version 10.1.0 (Oxford Instruments).

### DNA extraction and quantitative PCR (qPCR)

DNA was extracted as described earlier using a QIAamp DNA mini kit (Qiagen)^30, 36^. Bacterial density was quantified by qPCR reactions in a total 25 μl volume. Reactions contained 1x Platinum qPCR superMix (Invitrogen), 200 nm each of primers and probe, and 2.5 μl of purified DNA. The nucleotide sequence of primers and probe were published elsewhere^12^. No template controls (NTC) were run with each set of samples. qPCR reactions were carried out using a CFX96 Real-Time PCR Detection System (Bio-Rad). The following amplification parameters were utilized, 95°C for 2 min, followed by 40 cycles of 95°C for 15 s and 60°C for 1 min. qPCR standards using genomic DNA from reference strain TIGR4 (ATCC BAA-334) were run in parallel to construct a standard curve utilized to calculate genome equivalents (GenEq)/ml using the software Bio-Rad CFX manager. Standards DNA Preparation: Purified DNA was adjusted to a concentration of 1 ng/μl in TE buffer (10 mM Tris-HCl, 1 mM EDTA, pH 8.0) and immediately stored at -80°C until use. Standards for quantification were prepared within an hour before reactions by serially diluting a 1 ng/μl aliquot in TE buffer to final concentrations of 100 pg/μl, 10 pg/μl, 1 pg/μl, 100 fg/μl, 50 fg/μl, and 5 fg/μl of pneumococcal DNA. Given the 2.1608 Mb genome size of TIGR4^20^, these standards corresponded to 4.29×10^5^, 4.29×10^4^, 4.29×10^3^, 4.29×10^2^, 4.29×10^1^, 2.14×10^1^, or 2.14 GenEq, respectively. Standards prepared using this protocol consistently achieved an efficiency greater than 90% throughout the study (data not shown).

### Isolation and identification of *S. pneumoniae* strains

Nasopharyngeal specimens were thawed, vortexed for 15 s and 200 μl transferred to a 5 ml Todd-Hewitt broth (THY) containing 0.5% of yeast extract and 1 ml of rabbit serum (Gibco® by Life Technologies)^37^. This enriched culture was incubated for 6 h at 37°C in a 5% CO_2_ atmosphere and then inoculated onto BAP and incubated for 18-24 h at 37°C in a 5% CO_2_ atmosphere. Spn strains were isolated and identified using the optochin test (Remel) and bile solubility test as previously described^37^.

Spn strains were isolated and identified from lower respiratory tract specimens including sputum, endotracheal aspirates, bronchoalveolar lavage fluids, bronchial washings, protected brush specimens, and lung aspirates, using standard methodologies^38^. Briefly, after Gram staining, the primary media including 5% Columbia sheep blood, Chocolate, and MacConkey agars (BD BBL™) were inoculated. Thioglycolate broth was employed for bronchial brush samples. The media were incubated for 18 to 24 h at 35-37°C in 5% CO_2_, and cultures that remained negative after this period were re-incubated for another 24 h.

Pathogenic organisms were identified and susceptibility testing was performed only on significant growth, characterized by moderate to abundant colonies in the second or further quadrants of the plate, small numbers of a pathogen consistent with the predominant Gram-stain morphotype, and associated with inflammatory white blood cells, or colonies in the first quadrant if minimal or no other normal flora was present. Alpha-hemolytic, dry, or mucoid colonies resembling Spn strains were identified using Matrix Assisted Laser Desorption Ionization Time-of-Flight (VITEK® MS) or VITEK® 2 Gram-Positive identification card (GP), and purity was confirmed with the optochin disc. The antibiotic sensitivity was determined using VITEK® 2 Gram-Positive Susceptibility Cards, and the manual D-test.

### Statistical analysis

We performed one-way analysis of variance (ANOVA) followed by Dunnett’s multiple-comparison test when more than two groups were compared or Student’s t test to compare two groups, as indicated. All statistical analysis was performed using the software GraphPad Prism (version 8.3.1).

## Results

### *In vitro* assessment of fluorescently-labeled anti-Spn antibodies for detecting *S. pneumoniae* strains

We first evaluated the ability of fluorescently-labeled antibodies to detect specific Spn strains. Antibodies targeting vaccine serotypes, or serogroups, were labeled as detailed in Material and Methods. In addition to their specific reactivity against corresponding serotypes/serogroups, the labeled antibodies did not exhibit cross reactivity against a panel of other serotypes (not shown). For instance, the anti-S4-A488 (green), or anti-S19-A555 (red) antibody specifically stained Spn serotype 4 or 19F, respectively, within five min of exposure as evaluated by fluorescence microscopy (Fig. 1A and not shown). High-resolution micrographs using confocal microscopy were obtained from S19F strain 4924 growing on a glass substratum (Fig. 1B).

**Figure 1.**
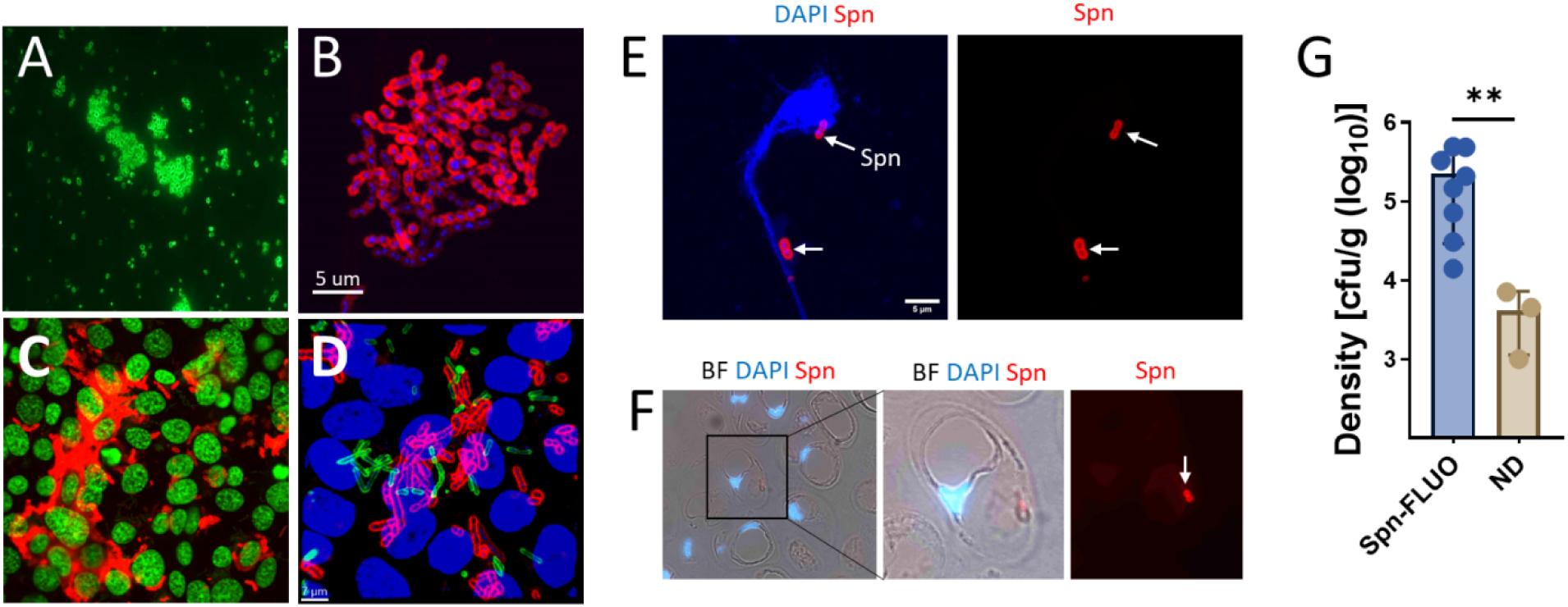
*In vitro* and *in vivo* detection of pneumococci with fluorescently-labelled antibodies. Strain TIGR4 (A) was mixed with anti S4-A488 (green) antibody and observed under a epifluorescence microscope within five minutes. (B) Spn serotype 19F strain 4924 was cultured for four h in a 8-well slide and pneumococci attached to the substratum were stained with an anti S19-A555 (red) antibody and the DNA was stained with DAPI. (C-D) Human pharyngeal cells grown to confluence were inoculated with (C) strain D39 or (D) a mixture of strain D39 and strain TIGR4 and incubated for four hours. D39 was stained with S2-A555 (red) and TIGR4 was stained with S4-A488. In panel C, DNA was stained with TO-PRO-3, while in panel D, it was stained with DAPI. (E-F) C57BL/6 mice (N=11) were intranasally inoculated with serotype 19F strain EF3030. After 48 h, mice were euthanized, and the nasal bone was removed. Nasopharyngeal tissue was collected, sectioned (∼5 µm), or homogenized. Nasopharyngeal homogenate (E) or tissue sections (F) were stained with DAPI and with an anti S19-A555 antibody. Arrows point out Spn. In panels B-F, micrographs were obtained by confocal microscopy, and the projection of z-stacks is shown. (G) Nasopharyngeal homogenates were diluted and plated to obtain the bacterial density. The density in the plot was grouped according to whether the samples yielded a positive reaction with Spn-FLUO or were not detected (ND). Student t test, \*\**p*=0.01.

We next evaluated the detection of Spn strains using fluorescently labeled antibodies in strains infecting *in vitro*-cultured human pharyngeal cells. Pharyngeal cells were infected for four h with strain D39 (serotype 2) or with a mixture of strains D39 and TIGR4 (serotype 4), and subsequently stained with anti-S2-A555 or with a combination of anti-S2-A555 and anti-S4-A488. Cell nuclei were also stained with a fluorescent dye. Spn strains were identified in cells infected with a single serotype (Fig. 1C). Additionally, each individual Spn strain was detected on pharyngeal cells infected with the D39-TIGR4 mixture (Fig. 1D).

### Detection of Spn vaccine strain serotype 19F in the mouse upper respiratory tract

A murine model of nasopharyngeal carriage was employed to assess the utility of fluorescent antibodies for detecting Spn strains. Eleven mice were intranasally inoculated with ∼10^5^ colony-forming units (CFU) of Spn strain EF3030, a serotype 19F strain commonly used in animal models of pneumococcal carriage^24, 39^. This inoculum was previously optimized to induce colonization levels comparable to those observed in the nasopharynx of children^33, 40^.

Two days post-infection, nasopharyngeal tissue from the upper respiratory tract was collected and stored in paraformaldehyde or homogenized. An aliquot of the nasopharyngeal homogenate was subjected to serial dilution and plating to quantify the pneumococcal burden. All mice were colonized with a median bacterial load of 7.2×10^4^ cfu/organ (interquartile range: 7.1×10^4^ - 3.3×10^5^) (Table 1). Subsequently, aliquots of the nasopharyngeal homogenates were subjected to pneumococcal detection assays. We employed an anti S19-A555 antibody, known to recognize strain serotype 19F EF3030^41^, and Spn-FLUO antibody raised against 92 pneumococcal serotypes, including serotype 19F. Both antibodies detected pneumococci in 72% of the specimens (8/11) (Fig. 1E and Table 1). Detection of pneumococci was not achieved in nasopharyngeal homogenates with a bacterial density ≤7.1×10^3^ cfu/organ (Fig. 1G and Table 1). Histological analysis of nasopharyngeal tissue revealed intracellular pneumococci within goblet cells indicating a good performance of antibody-based detection at the tissue level (Fig. 1F).

**Table 1.** Detection of pneumococci using Spn-FLUO in the upper respiratory tract of mice.

| Mouse nasopharynx | Density (cfu/organ) | Spn-FLUO* | S19-A555* |
| --- | --- | --- | --- |
| MN-1 | $7.1 \times 10^3$ | ND | ND |
| MN-2 | $3.27 \times 10^5$ | + | + |
| MN-3 | $4.93 \times 10^5$ | + | + |
| MN-4 | $1.4 \times 10^4$ | + | + |
| MN-5 | $1.47 \times 10^5$ | + | + |
| MN-6 | $4.83 \times 10^5$ | + | + |
| MN-7 | $1.0 \times 10^3$ | ND | ND |
| MN-8 | $4.5 \times 10^3$ | ND | ND |
| MN-9 | $7.2 \times 10^4$ | + | + |
| MN-10 | $3.07 \times 10^4$ | + | + |
| MN-11 | $2.09 \times 10^5$ | + | + |

\*ND=not detected.
+, up to five pneumococci per field in at least three fields of five observed using the 60x objective.

### Phenotypic and genotypic identification of *S. pneumoniae* in human nasopharyngeal specimens

For this initial proof-of-concept study, we selected fifty nasopharyngeal specimens from children from our previous studies^28, 30, 42^ and were divided into two groups (N=25 each) as detailed in Table 2. In one group, containing nasopharyngeal specimens listed as 1 through 25, Spn strains were isolated and identified using both traditional and genomic methods. In the other group (i.e., nasopharyngeal specimens 26 through 50) Spn cultures were negative.

**Table 2.**
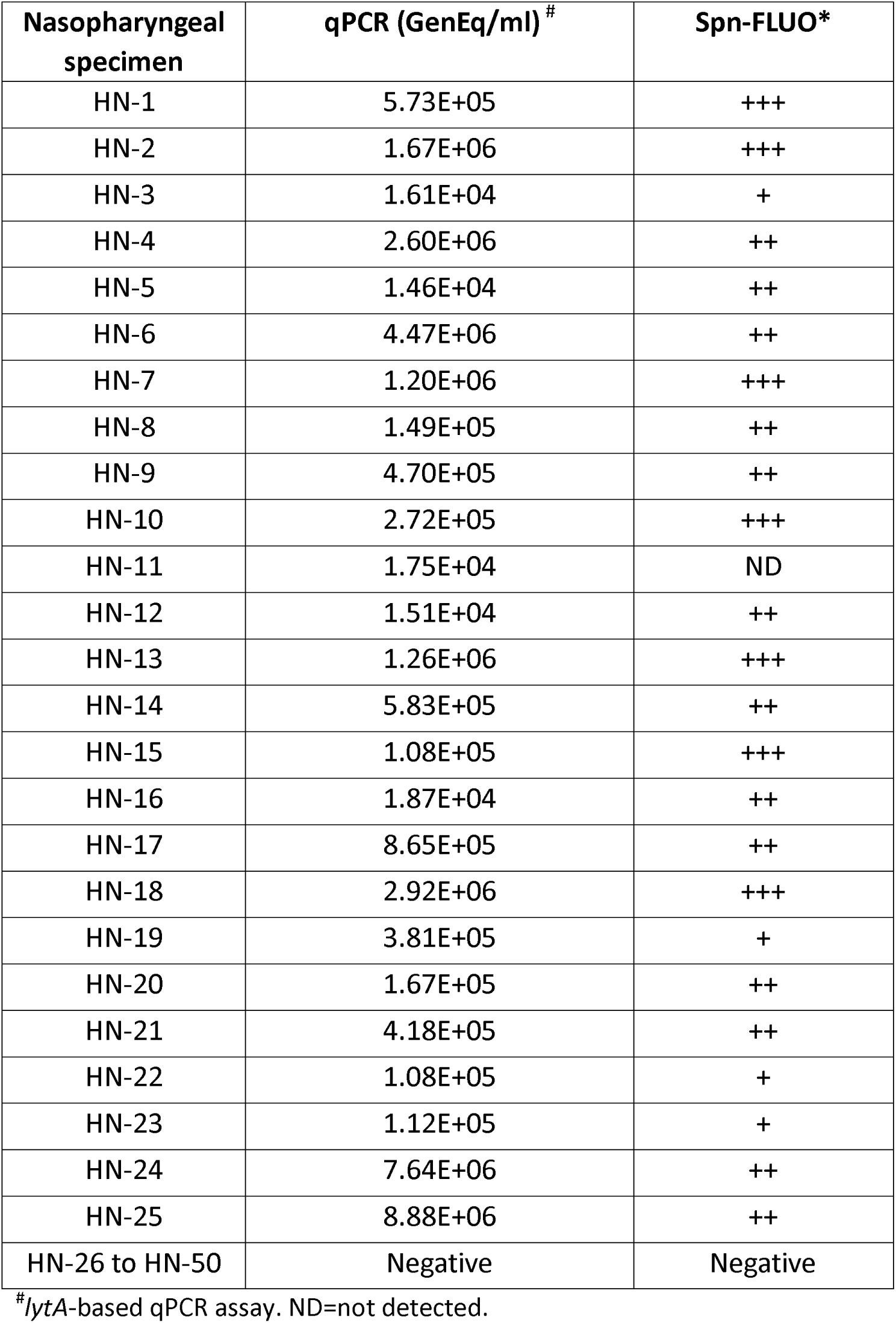
Detection of Spn in nasopharyngeal specimens from children with Spn-FLUO.

DNA was extracted from these nasopharyngeal samples and utilized as template in qPCR reactions targeting the species-specific *lytA* gene. Nasopharyngeal specimens 26 through 50 tested negative for Spn using *lytA*-PCR. As expected, *lytA*-PCR was positive for all nasopharyngeal specimens 1 to 25 (Table 2). The specimens were processed microscopically using fluorescently labeled antibodies, following the algorithm outlined in Figure 2A. Five distinct fields were examined within each preparation. The sensitivity of Spn-FLUO for detecting Spn strains in human nasopharyngeal specimens was 96% (24/25), while specificity was 100% as no Spn was detected in *lytA*-negative, culture-negative specimens. A typical microscopy finding from specimens containing abundant Spn (i.e., +++) is shown in Fig. 2C-D. The lowest bacterial density detected by fluorescent antibody staining was 1.61×10^4^ GenEq/ml (Table 2 and Fig. 2B).

**Figure 2.**
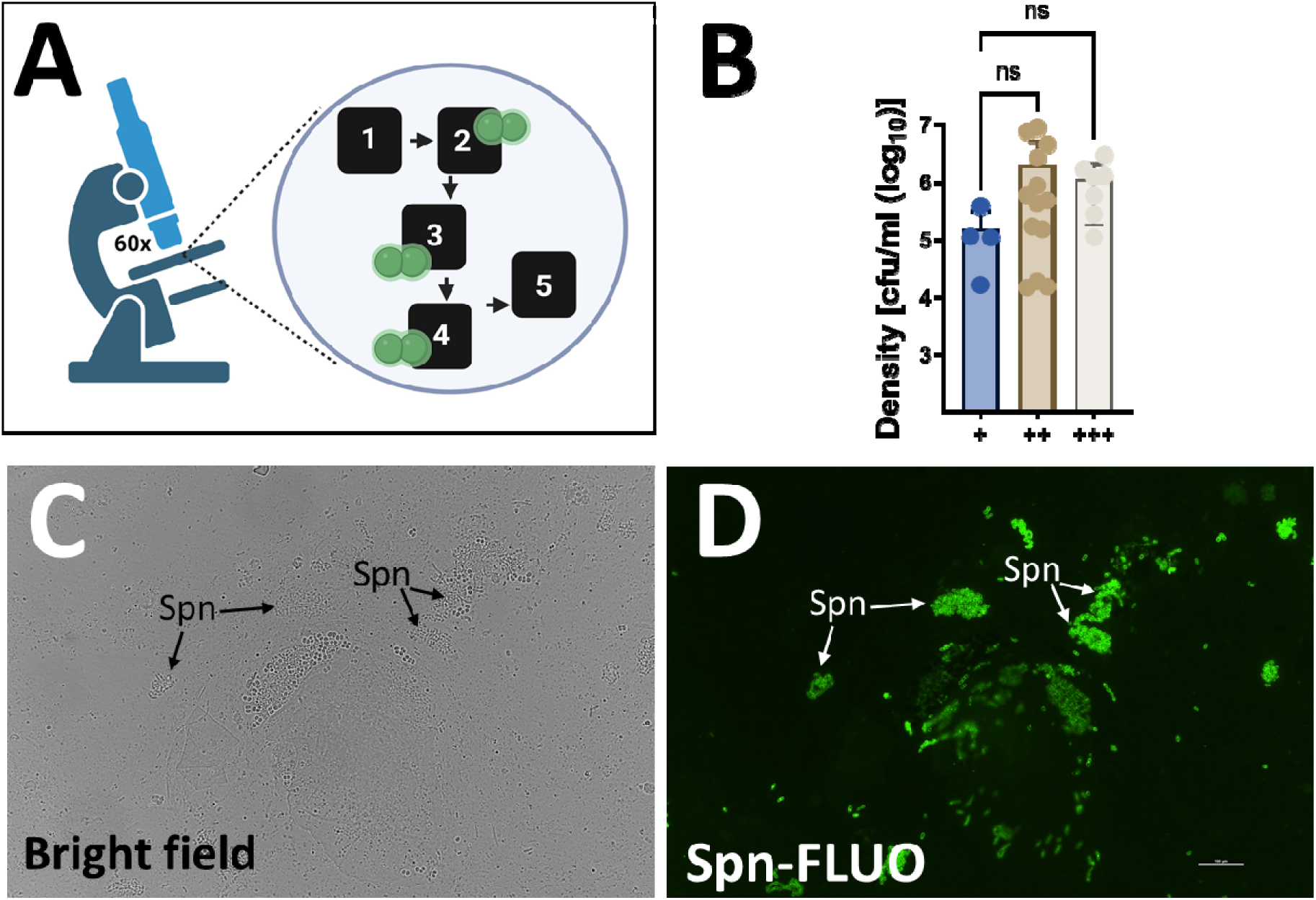
Detection of *S. pneumoniae* with Spn-FLUO in human specimens. (A) Detection algorithm for the identification of Spn in clinical specimens. Five fields were screened using the 60x objective, and a specimen was scored positive when ≥1 to <5 (+), ≥5 to <10 (++), or ≥10 (+++) pneumococci were observed in at least three fields. This algorithm was chosen to ensure the reliable detection of Spn while minimizing the risk of false positives. (B) The density of *S. pneumoniae* in nasopharyngeal specimens that yielded a positive reaction with Spn-FLUO was categorized depending on the semiquantitative algorithm. Data are presented as mean ± SD. Differences in bacterial density were determined using one-way ANOVA followed by Dunnett’s multiple comparisons test. ns=no significant difference. (C-D) Sputum specimen stained with Spn-FLUO and observed under (C) bright field and (D) epifluorescence in the green channel. Arrows point out Spn identified by Spn-FLUO.

### Detection and quantification of *Streptococcus pneumoniae* in mouse lung tissue during pneumonia

We next conducted a pre-clinical evaluation of Spn-FLUO for the detection of pneumococci in lung specimens using a murine model of pneumococcal pneumonia. Mice were infected with Spn strain TIGR4 and euthanized when moribund. Lungs were collected, homogenized, and the Spn bacterial load was determined by serial dilution and plating (Table 3). Lungs from mice infected with an isogenic capsule-deficient mutant (Δ*spxB*Δ*lctO*) that has a defect for colonization^43, 44^ served as controls. As expected, all animals infected with TIGR4 exhibited a high lung colonization density of 1.18×10^7^ (interquartile range: 2.37×10^6^ - 1.12×10^8^) colony-forming units (CFU)/g, whereas those infected with the isogenic mutant displayed low colonization levels (Fig. 3A). Spn was detected in all lung specimens from TIGR4-infected animals at varying levels of positivity. In contrast, mutant-infected animals exhibited significantly lower bacterial burdens, with <1×10^3^ cfu/g or no detectable bacteria (Fig. 3B, 3C and Table 3). Specimens with the highest density and positive Spn-FLUO detection were statistical different compared to those containing a lower burden of pneumococci (Fig. 3C). The highest positive reaction with Spn-FLUO was achieved in lungs from mice colonized with a median of 1.41×10^8^ CFU/g, which was statistically significant compared to groups with lower detection levels (Fig. 3B). Thus, Spn-FLUO demonstrated a strong correlation between fluorescence detection of pneumococci and bacterial load in a murine model of pneumococcal pneumonia, suggesting its potential as a rapid and sensitive diagnostic tool.

**Figure 3.**
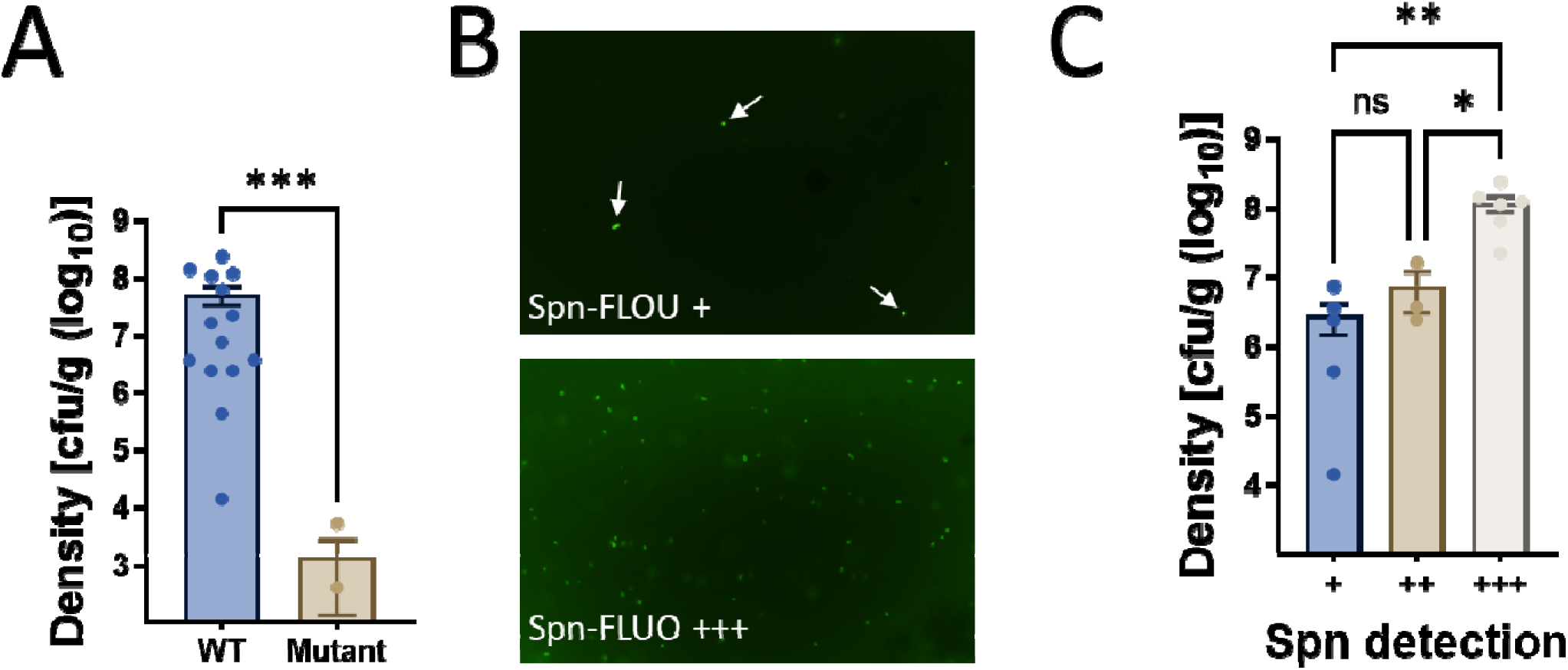
Detection of *S. pneumoniae* with Spn-FLUO in mouse lung specimens. Balb/c mice were intranasally inoculated with strain TIGR4 or isogenic mutant Δ*spxB*Δ*lctO* and euthanized when deemed moribund, or after 96 h post-infection. Lungs were collected and homogenized. (A) Lung homogenates were diluted and plated to obtain the bacterial density. (B) Aliquots of lung homogenate were stained with Spn-FLUO and scored for the presence of pneumococci. Micrographs were obtained with a fluorescence microscope in the green channel. The top panel shows a representative specimen scored “+” with arrows pointing out *S. pneumoniae*, while the bottom panel shows a representative specimen that scored “+++”. (C) The density in the graphic was grouped according to the semiquantitative detection of *S. pneumoniae* with Spn-FLUO. Statistical significance of differences in bacterial density were determined using Student’s t-test (A) or one-way ANOVA followed by Dunnett’s multiple comparisons test (C). *p=0.027, **p=.0088, ***p=0.0007.

**Table 3.**
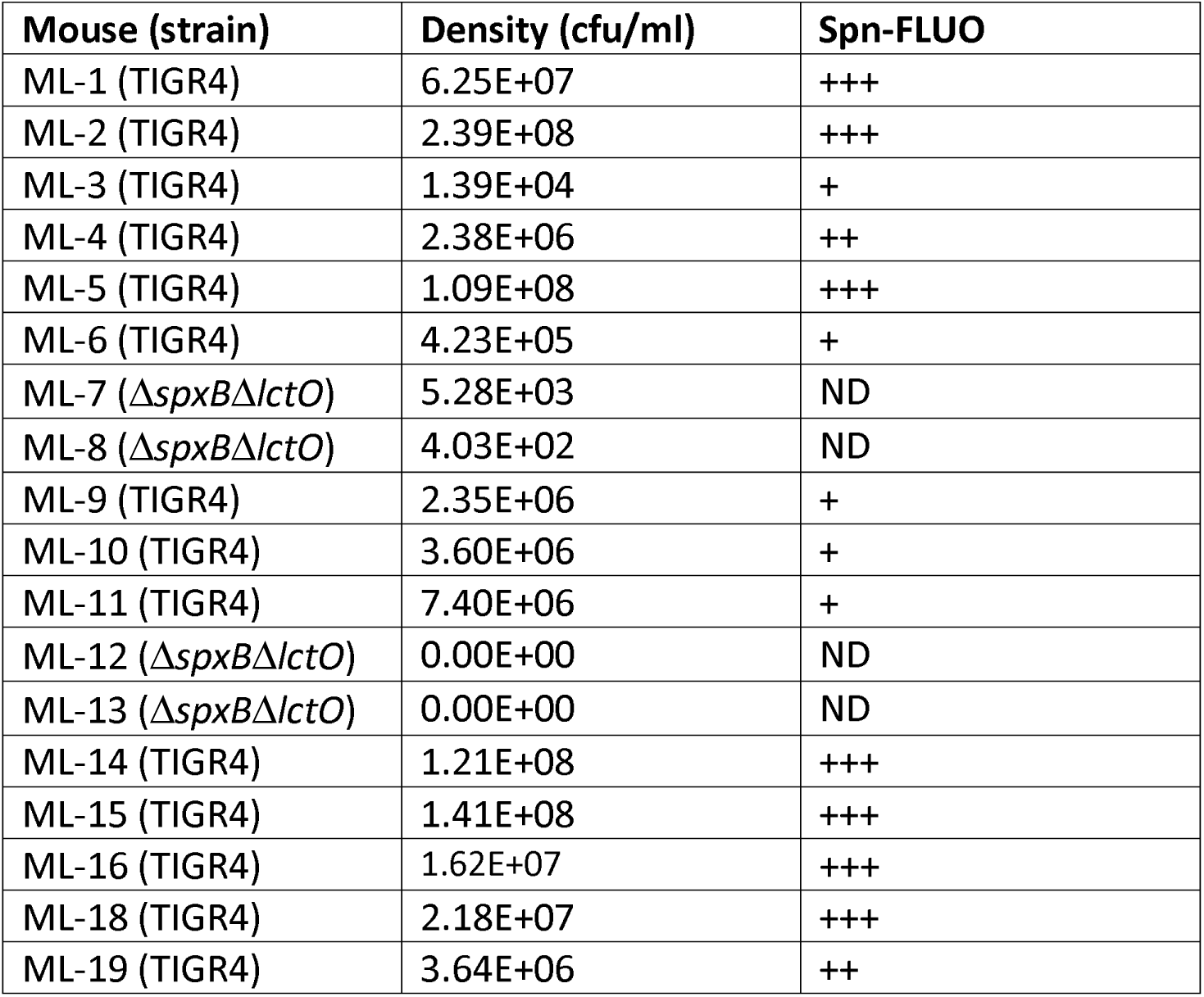
Detection of pneumococci using Spn-FLUO in the lung of mice.

### Detection of pneumococcal strains using Spn-FLUO in clinical specimens from patients with pneumococcal pneumonia

We subsequently evaluated the sensitivity of Spn-FLUO antibodies in detecting pneumococcal strains within human specimens from patients with pneumococcal pneumonia. To assess this, we analyzed N=27 clinical specimens obtained from patients with microbiologically confirmed pneumococcal pneumonia, each yielding a pneumococcal isolate. Specimens comprised sputum and bronchioalveolar lavage fluid (Table 4). DNA was extracted from all 27 specimens, each of which yielded a positive *lytA*-based qPCR result, indicating varying bacterial loads expressed as genome equivalents per milliliter (GE/ml). Consistently, all specimens were also positive for *piaB* (data not shown).

**Table 4.** Detection of Spn in culture-positive specimen from patients with pneumococcal pneumonia using Spn-FLUO.

| Specimen | ID | Specimen | qPCR<br>(GE/ml)* | Spn-FLUO |
| --- | --- | --- | --- | --- |
| HL1 | 082W | Sputum | $3.25 \times 10^6$ | + |
| HL2 | U019 | BAL & (endotracheal Tube) | $5.45 \times 10^7$ | +++ |
| HL3 | U0134 | Sputum | $6.16 \times 10^5$ | + |
| HL4 | U146 | Sputum | $5.10 \times 10^5$ | + |
| HL5 | U265 | Sputum | $3.63 \times 10^3$ | + |
| HL6 | U2275 | BAL (endotracheal Tube) | ND | +++ |
| HL7 | U6831 | BAL (endotracheal Tube) | $7.29 \times 10^9$ | + |
| HL8 | U077 | Sputum | $2.73 \times 10^5$ | + |
| HL9 | U097 | Sputum | $1.84 \times 10^5$ | + |
| HL10 | U260 | Sputum | $1.81 \times 10^6$ | + |
| HL11 | U227.46 | BAL (endotracheal Tube) | $4.85 \times 10^8$ | +++ |
| HL12 | U080 | Sputum | $1.10 \times 10^7$ | + |
| HL13 | U081 | Sputum | $1.31 \times 10^7$ | ++++ |
| HL14 | U081-W | Sputum | $7.54 \times 10^4$ | + |
| HL15 | U220 | Sputum | $2.60 \times 10^5$ | + |
| HL16 | U766 | Sputum | $6.36 \times 10^5$ | - |
| HL17 | U156 | Sputum | $7.56 \times 10^5$ | + |
| HL18 | U209 | Sputum | $2.67 \times 10^6$ | + |
| HL19 | U210 | Sputum | $5.32 \times 10^4$ | - |
| HL20 | U501 | Sputum | $3.95 \times 10^4$ | ++ |
| HL21 | UC1 | Sputum | $5.06 \times 10^7$ | ++ |
| HL22 | UC2 | Throat swab | $1.48 \times 10^5$ | ++ |
| HL23 | UC3 | Throat swab | $1.57 \times 10^4$ | + |
| HL24 | UC4 | Sputum | $4.57 \times 10^7$ | ++++ |
| HL25 | UC4 | Endotracheal aspirate | $1.13 \times 10^8$ | ++++ |
| HL26 | UC5 | BAL (endotracheal tube) | $3.84 \times 10^8$ | +++ |
| HL27 | UE1 | BAL (endotracheal tube) | $8.19 \times 10^6$ | ++ |

\*Genome equivalent/ml; & Bronchoalveolar lavage,

Spn-FLUO identified the pneumococcal etiology in 25/27 specimens, resulting in percent positive agreement of 92.6%. Specimens with high pneumococcal detection per field using Spn-FLUO contained a median bacterial load of 1.13×10^8^ GE/ml by *lytA*-based qPCR (interquartile range: 4.57×10^7^ - 4.85×10^8^) (Fig. 4A and Table 4). Specimens with low-moderate (i.e., scored +, and ++) detection of Spn per field had a median bacteria load of 5.10×10^5^ GE/ml (interquartile range: 1.11×10^5^ - 2.96×10^6^), while the specimen with the lowest GE/ml that yielded a positive reaction with the Spn-FLUO antibodies contained 3.63×10^3^ GE/ml of pneumococcal DNA (Fig. 4A).

**Figure 4.**
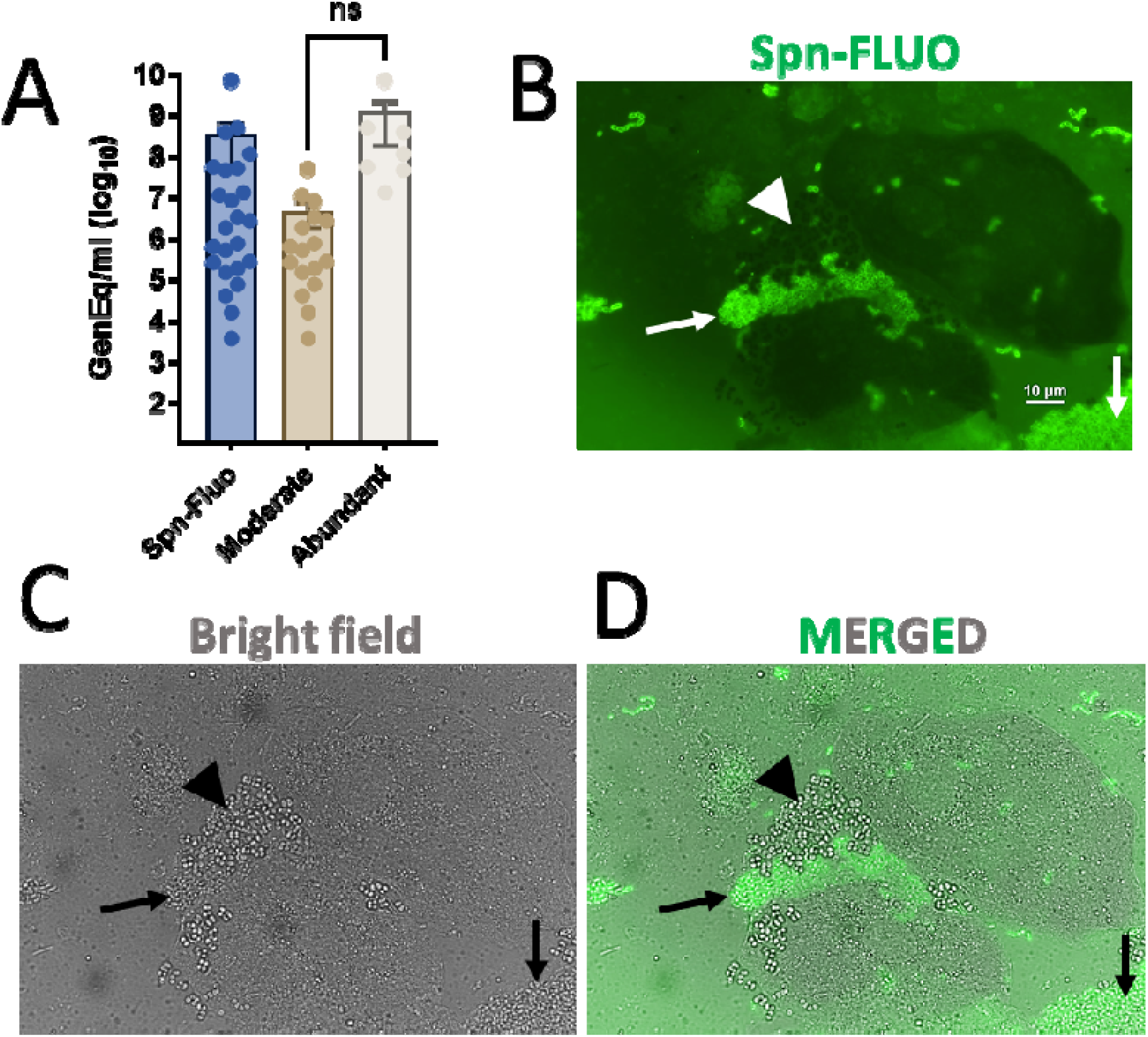
Detection of *S. pneumoniae* with Spn-FLUO in human sputum. (A) Aliquots of sputum were DNA extracted and used as templates in qPCR reactions targeting the *lytA* gene. Other aliquots were stained with Spn-FLUO, and the presence of *S. pneumoniae* was scored. The graphic shows the pneumococcal genome equivalent (GenEq)/ml in all samples, categorized as low (scored +), medium (scored ++), or high (scored +++). Statistical significance of differences in bacterial density was assessed using the Student’s t-test. ns=no significant difference. (B-D) Sputum specimens stained with Spn-FLUO and observed under (B) epifluorescence in the green channel or (C) bright field. Panel (D) shows the merged epifluorescence and bright field channels. Arrows point out *S. pneumoniae* identified by Spn-FLUO, while arrowheads signal staphylococcus-like microcolonies, which were not stained by Spn-FLUO but were observed in the bright field channels.

Representative images of Spn detected in sputum specimens are shown in Figure 4B-D. Fluorescence and bright-field microscopy revealed a heterogeneous mixture of epithelial cells, leukocytes, and bacteria resembling pneumococcal aggregates (Fig. 4B-D, arrows). While pneumococcal chains were also observed, these were less abundant than the bacterial aggregates. In a sputum specimen from a patient with pneumococcal pneumonia, additional bacteria were noted, although only pneumococci were stained by Spn-FLUO (Figure 4B-D, arrowhead). In summary, Spn-FLUO detected Spn strains in 92.6% of clinical specimens from patients with pneumococcal pneumonia within 30 min.

### Ultrastructural studies of *Streptococcus pneumoniae* in human sputum

Given the presence of abundant mucus and thick smears in some specimens, as well as the observation of large pneumococcal aggregates in unprocessed sputum, we performed high-resolution 3D confocal analysis of sputum specimens. Sputum was stained with Spn-FLUO, DAPI, and wheat germ agglutinin (WGA which binds to N-acetyl-D-glucosamine (GlcNAc) and sialic acid (SA) residues. GlcNAc/SA was utilized since sialic acid is a biomarker for mucin content^45^. A middle optical section of the XY, XZ (bottom panel), and YZ (side panel) focal planes revealed a large structure composed of aggregated pneumococci, isolated smaller aggregates, and pneumococcal chains (Fig. 5A and Video S1). An optical zoom revealed abundant DNA located adjacent to pneumococcal aggregates, suggesting the release from neighboring cells or cellular lysis (Fig. 5B). Orthogonal XZ and YZ sections demonstrated that the aggregates were embedded within the sputum (Fig. 5B, bottom panel). Pneumococci in closed proximity to cell nuclei, either in the process of capsule loss or already decapsulated (i.e., D-CPS), were observed in regions of the sputum resembling internalized bacteria (Fig. 5C).

**Figure 5.**
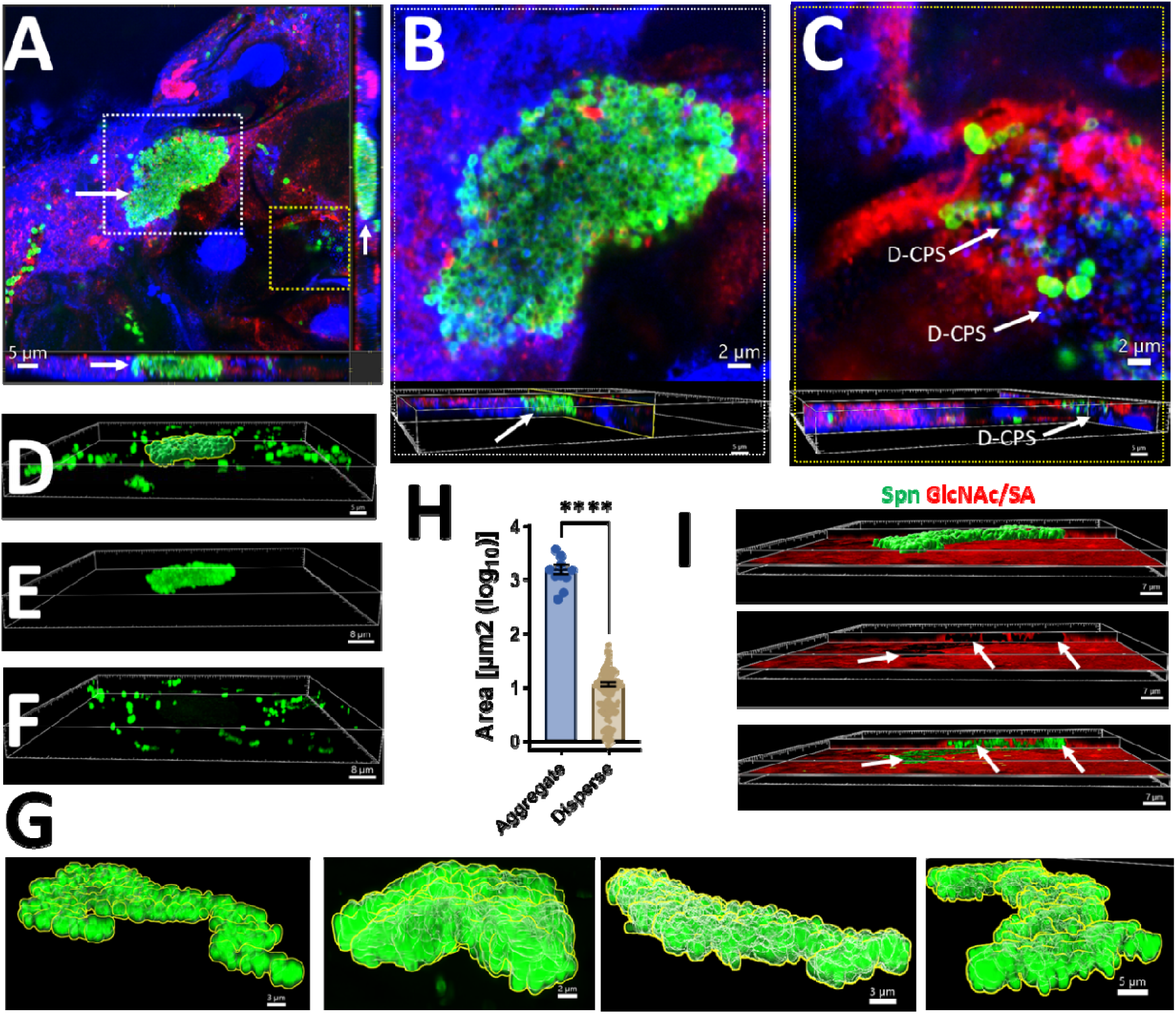
Ultrastructural analysis of *S. pneumoniae* in human sputum. Sputum specimens were stained with Spn-FLUO (green), wheat germ agglutinin (WGA) labeled with Alexa 555 (red), and DNA was stained with DAPI (blue). Preparations were analyzed by confocal microscopy. Panel (A) shows middle XY, XZ (bottom), and YZ (side) optical sections. The white inset indicates an optical zoom shown in panel (B), while the yellow inset indicates the region shown in panel (C). The bottom panels in (B and C) show orthogonal XZ and YZ sections. Decapsulated pneumococci (D-CPS) are indicated by the arrows. (D-F) A mask was created in the green channel (D) to analyze the 3D structure of the pneumococcal (E) aggregate or (F) dispersed pneumococci. (G) Representative 3D images of aggregated pneumococci across the sputum specimen, showcasing their size, shape, and spatial distribution within the sputum. (H) The area of those aggregate pneumococci and that of dispersed bacteria was plotted. Student’s t-test was performed to assess statistical significance, \*\*\*\**p*<0.0001. (I) Orthogonal XY and YZ sections were created using the green (Spn-FLUO) and the red channel (WGA). WGA binds acetyl glucosamine and sialic acid residues (GlcNAc/SA). The top panel shows the masked green channel representing the area of the pneumococcal aggregate. The pneumococcal aggregate was removed from the middle panel to reveal the hollow spaces occupied by bacteria embedded in the sputum. The bottom panel shows the non-masked green channel and the red channel. All analyses were performed using the Imaris software.

We performed three-dimensional reconstruction of the capsule channel (green) from confocal z-stacks (Fig. 5D) and categorized pneumococci into two groups: those within aggregates (Fig. 5E) and those forming chains or dispersed within the preparation (Fig. 5F). Representative pneumococcal aggregates are shown in Fig. 5G. The area of these pneumococcal aggregates was significantly different compared to all other pneumococci (Fig. 5H). The median area of pneumococcal aggregates was 1336 µm^2^ (interquartile range: 959 – 2108 µm^2^) whereas the median area of pneumococcal chains and dispersed bacteria was 9.15 µm^2^ (interquartile range: 3.6 – 14.3 µm^2^) (Fig. 5G).

Further 3D analysis of orthogonal XY and XZ focal planes demonstrated that the GlcNAc/SA signal did not colocalize with pneumococcal aggregates but instead surrounded encapsulated pneumococci (Fig. 5I). Overall, the data revealed that pneumococcal aggregates embedded in sputum specimens are significantly larger than those in chain or dispersed forms, potentially providing a physical barrier shielding pneumococci from sputum components.

## Discussion

In this study, we demonstrated the feasibility of fluorescent antibody-based microscopy for the rapid diagnosis of pneumococcal pneumonia. The entire process, from sample collection to final diagnosis, was completed within 30 min. Prompt identification of the causative agent is crucial in severe pneumonia, often influencing patient outcomes^9^. Spn-FLUO offers a valuable alternative for rapid diagnosis, particularly in cases where mortality remains high despite low antibiotic resistance rates, as observed with serotype 3 strains in Spain^46^. The rapid identification of the pneumococcus in clinical specimens allows for timely adjustment of empirical therapy, which is particularly important in severe cases. Although this technology requires a fluorescence microscope, the objective nature of the fluorescent signal from Spn strains makes fluorescence antibody-based detection suitable for resource-limited regions, as DNA extraction, reaction mixtures, and real-time systems are not necessary, and minimal training is required. Furthermore, clinical trials recruiting patients with pneumococcal pneumonia can benefit from a swift decision on whether potential individuals with signs and symptoms of pneumococcal pneumonia can be included in the trial.

Despite significant advancements in technology for the detection of pathogens causing pneumonia, pneumococcal pneumonia diagnosis remains challenging^9^. The culture of lower respiratory tract specimens, while considered the gold standard, is often associated with delays and low sensitivity. Multiplexed PCR panels have demonstrated improved sensitivity to detect pathogens not recovered by culture, but they require substantial resources, specialized equipment, and highly trained personnel, with turnaround times of several hours^9, 10, 47^. Antigen detection assays, such as those targeting the pneumococcal capsule, offer the advantage of rapid turnaround time. However, their specificity to a single pathogen represents a limitation. Nonetheless, they can be a life-saving tool in critical situations.

To the best of our knowledge, no previous studies have investigated the ultrastructure of pneumococci in freshly expectorated human sputum from patients with pneumonia. The current study filled this gap by characterizing the unique structures formed by pneumococci in the sputum. These bacterial aggregates are large, with a median size of 1336 µm², and have not been observed invading the lungs of patients with pneumococcal pneumonia or in the lungs of mice infected with Spn strains. This population of pneumococci, which likely resides extracellularly on bronchi and bronchioles, may be formed to resist the innate and adaptive immune response. These aggregated extracellular pneumococci represent an important novel population that may be more resistant to antibiotics and antimicrobial peptides.

Our study has some limitations. Notably, Spn-FLUO exclusively recognizes encapsulated pneumococci, limiting its utility for detecting non-encapsulated strains. While the majority of isolates express capsule, this limitation could hinder the detection of other non-encapsulated strains^48^. Additionally, studies evaluating Spn-FLUO sensitivity in both mouse lung specimens and human lung specimens from patients with pneumococcal pneumonia involved specimens from where strains were isolated in culture, suggesting a high bacterial load, later confirmed by qPCR assays. We did not assess the specificity of Spn-FLUO against a panel of Spn-negative human sputum samples or healthy subjects. Instead, as a surrogate, we utilized human nasopharyngeal specimens with a *lytA*-negative reaction and lung specimens from mice infected with a strain with a colonization defect and low capsule production. In both cases, specificity was 100%.

In summary, we have characterized a rapid method for detecting the pneumococcus in clinical specimens from patients with pneumonia. Using the fluorescent antibodies developed in this study, we have provided novel ultrastructural information regarding a previously uncharacterized population of pneumococcal aggregates. This study serves a dual purpose. First, antibody-based detection of pneumococci can be further developed to guide empirical therapy while awaiting culture results. Second, it fosters novel research into pneumococcal aggregates, which may serve as reservoirs for the pneumococcus.

## Data Availability

All data produced in the present work are contained in the manuscript

## List of abbreviations

Spn: *Streptococcus pneumoniae*
Spn-FLUO: Fluorescent antibodies against the pneumococcal capsule
qPCR: Quantitative polymerase chain reaction
LOD: Limit of Detection
GenEq/ml: Genome equivalents/milliliter
µm^2^: Square micrometer
min: Minutes
CAP: Community-acquired pneumonia
CO_2_: Carbon dioxide
O_2_: Oxygen
h: Hours
(m)PCR: Multiplex polymerase chain reaction
CFU: Colony Forming Units
CFU/g: Colony forming units/gram
WGA: Wheat Germ Agglutinin
GlcNAc: N-acetyl-D-glucosamine
SA: Sialic acid
BAP: Blood agar plates
THY: Todd Hewitt broth containing yeast extract
Str: Streptomycin
Tmp: Trimethoprim
Tet: Tetracycline
Ery: Erythromycin
µg/ml: Microgram/milliliter
PBS: Phosphate Buffered Saline
MEM: Minimum Essential Medium
FBS: Fetal Bovine Serum
PFA: Paraformaldehyde
BSA: Bovine Serum Albumin
µM: Micromole
IACUC: Institutional Animal Care and Use Committee
AWA: National Science Foundation Animal Welfare Act
IRB: Institutional Review Board
UMMC: University of Mississippi Medical Center
WHO: World Health Organization
NP: Nasopharyngeal
µl: Microliter
ng/ µl: Nanogram/microliter
Mb: Megabase
s: Seconds

## Declarations

### Ethics approval

The research project “Mechanistic Studies of Broadly Reactive Antibodies for the Treatment, Diagnosis, and Prevention of Pneumococcal Infection” has been reviewed by the Institutional Review Board (IRB) of the University of Mississippi Medical Center (Federalwide Assurance FWA#00003630) and has been determined to be exempt from formal review. The IRB concluded that the project meets the criteria for exemption as outlined in the U. S. Department of Health and Human Services Regulations for the Protection of Human Subjects, 45 CFR 46.102(l) because this project does not research involving human subjects. As such, no additional ethical review was required.

All experiments involving animals were performed with prior approval of and in accordance with protocol 1584 which was reviewed and approved by the UMMC Animal Care Committee. UMMC laboratory animal facilities have been fully accredited by the American Association for Accreditation of Laboratory Animal Care. Procedures were performed according to the institutional policies, Animal Welfare Act, NIH guidelines, and American Veterinary Medical Association guidelines on euthanasia.

### Consent to participate

N/A

### Consent for publication

N/A

### Availability of data and materials

All data generated or analyzed during this study are included in this published article (and its supplementary information files) and are available from the corresponding author on reasonable request. The materials such as software, protocols or reagents were specified in the Material and Methods section.

### Authors’ contributions

A.G.J.V.- Formal analysis, Investigation, Supervision, Methodology, Project administration, Writing - review & editing.

M.F. - Formal analysis, Investigation.

M.C.- Investigation.

I.J.F.- Investigation.

P.S. - Investigation.

P.V.- Investigation.

C.F. L.- Investigation, Methodology, Writing- review & editing.

C.G. - Investigation, Methodology, Writing - review & editing.

W.D. - Investigation.

J.E.V. -Conceptualization, Data curation, Formal analysis, Funding acquisition, Investigation, Writing - original draft, Writing - review & editing.

All authors read and approved the final manuscript.

## Acknowledgements

We thank Dr. Babek Alibayov from the University of Mississippi Medical Center (UMMC) for their assistance with labeling aliquots of Spn-FLUO, and Dr. Kenichi Takeshita, and Dr. Antonino Baez, also from UMMC, for their feedback on the manuscript. The authors also acknowledge the assistance of Dr. Fuminori Sakai from Emory University Rollins School of Public Health, currently at Pfizer Japan, for performing some qPCR assays.

## References

1. Wahl, B. et al. Burden of Streptococcus pneumoniae and Haemophilus influenzae type b disease in children in the era of conjugate vaccines: global, regional, and national estimates for 2000-15. Lancet Glob Health 6, e744–e757 (2018).

2. Weiser, J.N., Ferreira, D.M. & Paton, J.C. Streptococcus pneumoniae: transmission, colonization and invasion. Nat Rev Microbiol 16, 355–367 (2018).

3. Musher, D.M. & Thorner, A.R. Community-acquired pneumonia. N Engl J Med 371, 1619–1628 (2014).

4. Ganaie, F. et al. A New Pneumococcal Capsule Type, 10D, is the 100th Serotype and Has a Large cps Fragment from an Oral Streptococcus. mBio 11 (2020).

5. Brissac, T. et al. Capsule Promotes Intracellular Survival and Vascular Endothelial Cell Translocation during Invasive Pneumococcal Disease. mBio 12, e0251621 (2021).

6. Hyams, C., Camberlein, E., Cohen, J.M., Bax, K. & Brown, J.S. The Streptococcus pneumoniae capsule inhibits complement activity and neutrophil phagocytosis by multiple mechanisms. Infect Immun 78, 704–715 (2010).

7. Kim, J.O. et al. Relationship between cell surface carbohydrates and intrastrain variation on opsonophagocytosis of Streptococcus pneumoniae. Infect Immun 67, 2327–2333 (1999).

8. Miller, J.M. et al. A Guide to Utilization of the Microbiology Laboratory for Diagnosis of Infectious Diseases: 2018 Update by the Infectious Diseases Society of America and the American Society for Microbiology. Clin Infect Dis 67, e1–e94 (2018).

9. Pletz, M.W. et al. Unmet needs in pneumonia research: a comprehensive approach by the CAPNETZ study group. Respir Res 23, 239 (2022).

10. Walker, A.M., Timbrook, T.T., Hommel, B. & Prinzi, A.M. Breaking Boundaries in Pneumonia Diagnostics: Transitioning from Tradition to Molecular Frontiers with Multiplex PCR. Diagnostics (Basel) 14 (2024).

11. Marimuthu, S., Damiano, R.B. & Wolf, L.A. Performance Characteristics of a Real-Time PCR Assay for Direct Detection of Streptococcus pneumoniae in Clinical Specimens. J Mol Diagn 26, 552–562 (2024).

12. Carvalho Mda, G., et al. Evaluation and improvement of real-time PCR assays targeting lytA, ply, and psaA genes for detection of pneumococcal DNA. J Clin Microbiol 45, 2460–2466 (2007).

13. Wyllie, A.L. et al. Streptococcus pneumoniae in saliva of Dutch primary school children. PLoS One 9, e102045 (2014).

14. Tavares, D.A. et al. Identification of Streptococcus pneumoniae by a real-time PCR assay targeting SP2020. Sci Rep 9, 3285 (2019).

15. Shkodenko, L.A., Mohamed, A.A., Ateiah, M., Rubel, M.S. & Koshel, E.I. A DAMP-Based Assay for Rapid and Affordable Diagnosis of Bacterial Meningitis Agents: Haemophilus influenzae, Neisseria meningitidis, and Streptococcus pneumoniae. Int J Mol Sci 25 (2024).

16. Gotts, J.E. et al. Clinically relevant model of pneumococcal pneumonia, ARDS, and nonpulmonary organ dysfunction in mice. Am J Physiol Lung Cell Mol Physiol 317, L717–L736 (2019).

17. Lee, J.S., Giesler, D.L., Gellad, W.F. & Fine, M.J. Antibiotic Therapy for Adults Hospitalized With Community-Acquired Pneumonia: A Systematic Review. JAMA 315, 593–602 (2016).

18. Wu, X. et al. Effect of pneumococcal conjugate vaccine availability on Streptococcus pneumoniae infections and genetic recombination in Zhejiang, China from 2009 to 2019. Emerg Microbes Infect 11, 606–615 (2022).

19. Viteri-Davila, C. et al. The Crisis of Macrolide Resistance in Pneumococci in Latin America. Am J Trop Med Hyg (2024).

20. Tettelin, H. et al. Complete genome sequence of a virulent isolate of Streptococcus pneumoniae. Science 293, 498–506 (2001).

21. Wu, X. et al. Interaction between Streptococcus pneumoniae and Staphylococcus aureus Generates (.)OH Radicals That Rapidly Kill Staphylococcus aureus Strains. J Bacteriol 201 (2019).

22. McDevitt, E., et al. Hydrogen Peroxide Production by Streptococcus pneumoniae Results in Alpha-hemolysis by Oxidation of Oxy-hemoglobin to Met-hemoglobin. mSphere 5 (2020).

23. Lanie, J.A. et al. Genome sequence of Avery’s virulent serotype 2 strain D39 of Streptococcus pneumoniae and comparison with that of unencapsulated laboratory strain R6. J Bacteriol 189, 38–51 (2007).

24. Junges, R., Maienschein-Cline, M., Morrison, D.A. & Petersen, F.C. Complete Genome Sequence of Streptococcus pneumoniae Serotype 19F Strain EF3030. Microbiol Resour Announc 8 (2019).

25. Wu, X. et al. Competitive Dominance within Biofilm Consortia Regulates the Relative Distribution of Pneumococcal Nasopharyngeal Density. Appl Environ Microbiol 83 (2017).

26. Wu, X. et al. Ultrastructural, metabolic and genetic characteristics of determinants facilitating the acquisition of macrolide resistance by Streptococcus pneumoniae. Drug Resist Updat 77, 101138 (2024).

27. Alibayov, B. et al. Oxidative Reactions Catalyzed by Hydrogen Peroxide Produced by Streptococcus pneumoniae and Other Streptococci Cause the Release and Degradation of Heme from Hemoglobin. Infect Immun 90, e0047122 (2022).

28. Sakai, F., Sonaty, G., Watson, D., Klugman, K.P. & Vidal, J.E. Development and characterization of a synthetic DNA, NUversa, to be used as a standard in quantitative polymerase chain reactions for molecular pneumococcal serotyping. FEMS Microbiol Lett 364 (2017).

29. Pholwat, S., Sakai, F., Turner, P., Vidal, J.E. & Houpt, E.R. Development of a TaqMan Array Card for Pneumococcal Serotyping on Isolates and Nasopharyngeal Samples. J Clin Microbiol 54, 1842–1850 (2016).

30. Chien, Y.W. et al. Density interactions among Streptococcus pneumoniae, Haemophilus influenzae and Staphylococcus aureus in the nasopharynx of young Peruvian children. Pediatr Infect Dis J 32, 72–77 (2013).

31. Grijalva, C.G. et al. Cohort profile: The study of respiratory pathogens in Andean children. Int J Epidemiol 43, 1021–1030 (2014).

32. Howard, L.M. et al. Association between nasopharyngeal colonization with multiple pneumococcal serotypes and total pneumococcal colonization density in young Peruvian children. Int J Infect Dis 134, 248–255 (2023).

33. Nelson, K.N. et al. Dynamics of Colonization of Streptococcus pneumoniae Strains in Healthy Peruvian Children. Open Forum Infect Dis 5, ofy039 (2018).

34. Satzke, C. et al. Standard method for detecting upper respiratory carriage of Streptococcus pneumoniae: updated recommendations from the World Health Organization Pneumococcal Carriage Working Group. Vaccine 32, 165–179 (2013).

35. O’Brien, K.L. et al. Evaluation of a medium (STGG) for transport and optimal recovery of Streptococcus pneumoniae from nasopharyngeal secretions collected during field studies. J Clin Microbiol 39, 1021–1024 (2001).

36. Sakai, F., Talekar, S.J., Klugman, K.P. & Vidal, J.E. Expression of Virulence-Related Genes in the Nasopharynx of Healthy Children. PloS one 8, e67147 (2013).

37. Centers for Desease Control and Prevention & (CDC). Streptococcus pneumoniae carriage study protocol - nasopharyngeal (NP) swab processing.; 2010.

38. Leber, A.L. Clinical microbiology procedures handbook, 4th edition. edn. ASM Press: Washington, DC, 2016.

39. Scott, E.J., 2nd, Luke-Marshall, N.R., Campagnari, A.A. & Dyer, D.W. Draft Genome Sequence of Pediatric Otitis Media Isolate Streptococcus pneumoniae Strain EF3030, Which Forms In Vitro Biofilms That Closely Mimic In Vivo Biofilms. Microbiol Resour Announc 8 (2019).

40. Vidal, J.E. et al. Prophylactic inhibition of colonization by Streptococcus pneumoniae with the secondary bile acid metabolite deoxycholic acid. Infect Immun, IAI0046321 (2021).

41. Vidal, J.E. et al. Prophylactic Inhibition of Colonization by Streptococcus pneumoniae with the Secondary Bile Acid Metabolite Deoxycholic Acid. Infect Immun 89, e0046321 (2021).

42. Hanke, C.R. et al. Bacterial Density, Serotype Distribution and Antibiotic Resistance of Pneumococcal Strains from the Nasopharynx of Peruvian Children Before and After Pneumococcal Conjugate Vaccine 7. Pediatr Infect Dis J 35, 432–439 (2016).

43. Echlin, H., Frank, M., Rock, C. & Rosch, J.W. Role of the pyruvate metabolic network on carbohydrate metabolism and virulence in Streptococcus pneumoniae. Mol Microbiol 114, 536–552 (2020).

44. Alibayov, B., et al. Oxidation of hemoglobin in the lung parenchyma facilitates the differentiation of pneumococci into encapsulated bacteria. bioRxiv (2023).

45. Esther, C.R., Jr., et al. Sialic acid-to-urea ratio as a measure of airway surface hydration. Am J Physiol Lung Cell Mol Physiol 312, L398–L404 (2017).

46. Calvo-Silveria, S. et al. Evolution of invasive pneumococcal disease by serotype 3 in adults: a Spanish three-decade retrospective study. Lancet Reg Health Eur 41, 100913 (2024).

47. Riccobono, E., Bussini, L., Giannella, M., Viale, P. & Rossolini, G.M. Rapid diagnostic tests in the management of pneumonia. Expert Rev Mol Diagn 22, 49–60 (2022).

48. Keller, L.E., Robinson, D.A. & McDaniel, L.S. Nonencapsulated Streptococcus pneumoniae: Emergence and Pathogenesis. mBio 7, e01792 (2016).

